# Tracking Cryptic SARS-CoV-2 Lineages Detected in NYC Wastewater

**DOI:** 10.1101/2021.07.26.21261142

**Authors:** Davida S. Smyth, Monica Trujillo, Devon A. Gregory, Kristen Cheung, Anna Gao, Maddie Graham, Yue Guan, Caitlyn Guldenpfennig, Irene Hoxie, Sherin Kannoly, Nanami Kubota, Terri D. Lyddon, Michelle Markman, Clayton Rushford, Kaung Myat San, Geena Sompanya, Fabrizio Spagnolo, Reinier Suarez, Emma Teixeiro, Mark Daniels, Marc C. Johnson, John J. Dennehy

## Abstract

Tracking SARS-CoV-2 genetic diversity is strongly indicated because diversifying selection may lead to the emergence of novel variants resistant to naturally acquired or vaccine-induced immunity. To monitor New York City (NYC) for the presence of novel variants, we amplified regions of the SARS-CoV-2 Spike protein gene from RNA acquired from all 14 NYC wastewater treatment plants (WWTPs) and ascertained the diversity of lineages from these samples using high throughput sequencing. Here we report the detection and increasing frequencies of novel SARS-CoV-2 lineages not recognized in GISAID’s EpiCoV database. These lineages contain mutations rarely observed in clinical samples, including Q493K, Q498Y, H519N and T572N. Many of these mutations were found to expand the tropism of SARS-CoV-2 pseudoviruses by allowing infection of cells expressing the human, mouse, or rat ACE2 receptor. In addition, pseudoviruses containing the Spike amino acid sequence of these lineages were found to be resistant to many different classes of receptor binding domain (RBD) binding neutralizing monoclonal antibodies. We offer several hypotheses for the anomalous presence of these mutations, including the possibility of a non-human animal reservoir. Although wastewater sampling cannot provide direct inference of SARS-CoV-2 clinical sequences, our research revealed several lineages that could be relevant to public health and they would not have been discovered if not for wastewater surveillance.

## Main

SARS-CoV-2 is shed in feces and can be detected in wastewater correlating to caseloads in sewersheds^1,2^. Since January of 2021, we sequenced SARS-CoV-2 RNA isolated from all 14 NYC WWTPs approximately twice per month^3^. Our targeted sequencing strategy entailed iSeq 100 and MiSeq sequencing of PCR-amplified regions of the SARS-CoV-2 Spike protein gene, particularly the receptor binding domain (RBD) (Fig. 1A), spanning amino acid residues 434 to 505 for iSeq amplicons and 412 to 579 for MiSeq amplicons. These regions contain loci that are significant in SARS-CoV-2 receptor tropism and immune evasion, and contain multiple polymorphisms found in many variants of concern (VOC)^4,5^. Our analysis pipeline, which uses the tool SAM Refiner, allowed us to determine the frequency of each polymorphism and more importantly, elucidate which polymorphisms were derived from the same RNA sequence^6^.

**Figure 1.**
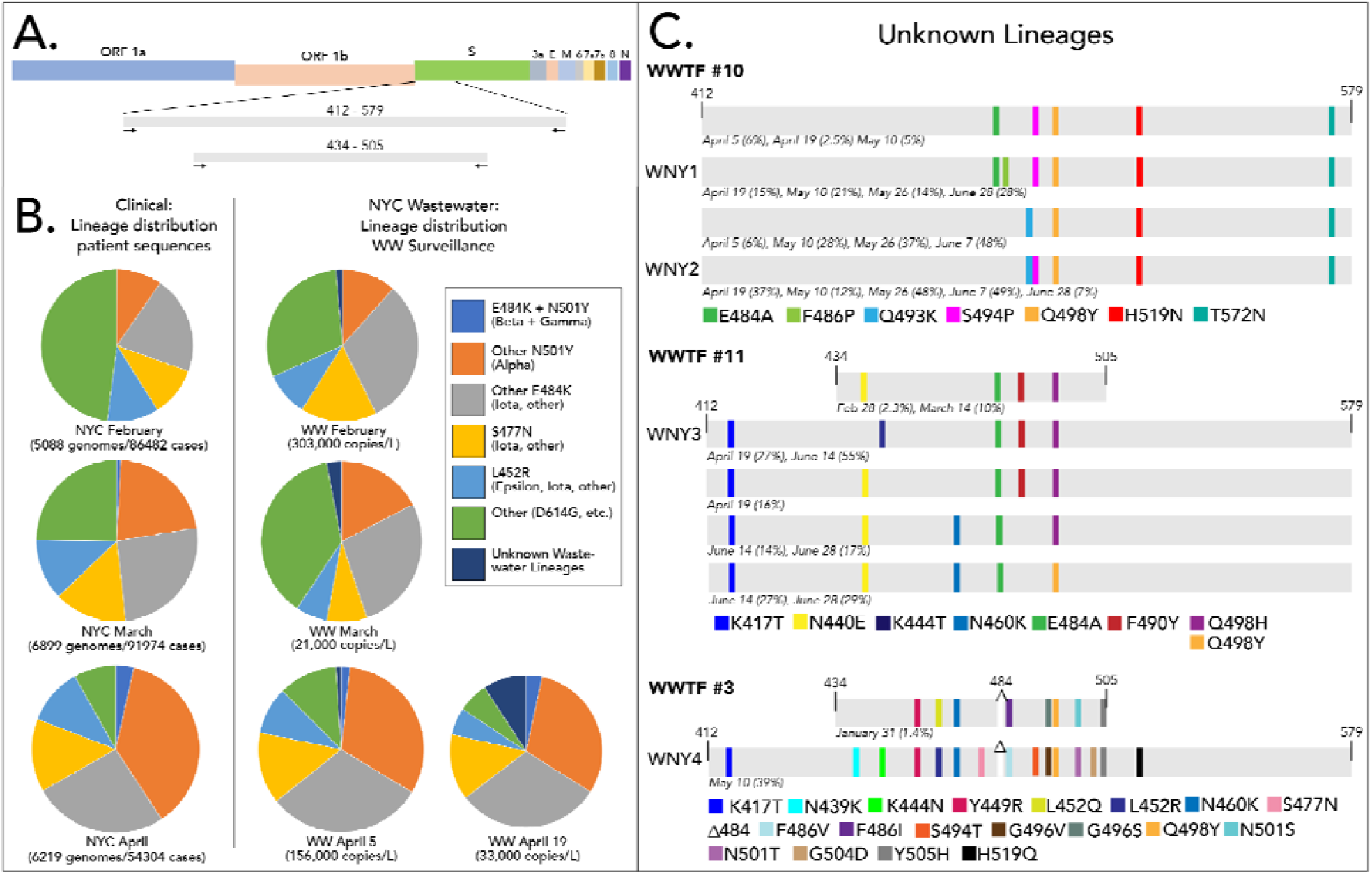
Novel SARS-CoV-2 lineages from Wastewater. A) Schematic of SARS-CoV-2 and the amplification locations. B) Distribution of SARS-COV-2 variants based on patient sequences and wastewater surveillance. C) Novel lineages detected. Schematic highlights shared sequences. Percentages indicate the percent of the sequences from each date that contained the indicated polymorphisms. Some sequences have additional polymorphisms not listed.

### Identification of Novel Sewershed-Specific Lineages

Using this approach, we were able to classify suites of mutations found in the RBD amplicons as consistent with Pango lineages B.1.1.7 (Alpha), B.1.351 (Beta), B.1.427/429 (Epsilon), B.1.526 (Iota), B.1.617 (Delta and Kappa) and P.1 (Gamma). Importantly, the distributions and trends in viral lineages from wastewater were consistent with patient derived sequences from NYC (Fig. 1B)(Supplemental Table 1). For example, between February and April, wastewater surveillance and patient sequencing both revealed a notable increase in sequences assigned to the Alpha lineage and a corresponding decrease in sequences that did not belong to any of the VOC lineages.

In addition to well-recognized lineages, three WWTPs, 3, 10, and 11, contained lineages with consistent, but not static, constellations of polymorphisms detected over several months that did not match lineages reported in the GISAID EpiCoV database (https://www.gisaid.org/)(Fig. 1C). Four of these lineages, designated WNY1, WNY2, WNY3, and WNY4, were selected for further study. Each of these lineages contained at least five polymorphisms; the most divergent was WNY4, which contained 16 amino acid changes in its RBD including a deletion at position 484.

Interestingly, all four novel lineages contained a polymorphism at position 498 (Q498H or Q498Y). As of July 26, 2021, there were only three US SARS-CoV-2 sequences in GISAID that contained the polymorphism Q498H, and none that contained Q498Y. However, both of these polymorphisms have been associated with host range expansion of SARS-CoV-2 into rodents^7–9^, which are generally resistant to the parent SARS-CoV-2 lineage^10–12^. Notably, as the concentration of SARS-CoV-2 genetic material from NYC wastewater decreased along with the decrease in COVID patients, the fraction of the total sequences from these unknown lineages has proportionally increased. By May and June, these lineages often represented the majority of sequences recovered from some treatment facilities (Fig. 1C). By May when cases were dramatically dropping, several of the NYC sewersheds did not contain high enough concentrations of SARS-COV-2 RNA for analysis, which prevented further determination of city-wide variant distributions from wastewater.

As an external confirmation of our findings, we analyzed raw reads uploaded to NCBI’s Sequence Read Archive (SRA) from nearly 1,500 other wastewater samples globally spanning 2020 and 2021 up to July 21, 2021, including 37 samples from New York state. Of all the samples, only 4, all from NY state sewersheds, had sequences matching the novel lineages we described (SRR15128978, SRR15128983, SRR15202284, SRR15202285).

### Are Cryptic Lineages Derived from Unsampled COVID-19 Infections?

The existence of these lineages may point to COVID-19 infections of human patients that are not being sampled through standard clinical sequencing efforts. The frequency of weekly confirmed cases in NYC that were sequenced ranged from 2.6% on January 31, 2021 to 12.9% on June 12, 2021^13^. Nonetheless, not all cases were diagnosed and not all positive samples were sequenced. Therefore, the cryptic lineages may be derived from asymptomatic, vaccinated, immunosuppressed, pediatric, or chronically infected patients who are not being sampled in clinical settings. Infectious SARS-CoV-2 in such patients may linger in the gut after infections have resolved in the respiratory tract^14–22^.

Alternatively, these lineages may be derived from physically distinct locations in the body. That is, perhaps viruses of these lineages predominantly replicate in gut epithelial cells, and are not present in the nasopharynx such that standard swabbing techniques can recover sufficient quantities for sequencing. Finally, we speculate that perhaps these mutations are found in minority variants that are unreported in consensus sequences uploaded to EpiCoV and other databases. Several groups have identified evidence of within host quasispecies in NGS datasets^23,24^. In one case, as many as 68% of the samples contained evidence of quasispecies in several loci, 76% of which contained nonsynonymous mutations concentrated in the S and orf1a genes^23^. To address whether our variants were associated with within-host diversity, we checked for minority variants in the raw reads of sequencing runs performed on samples collected between January 2020 to July 2021 obtained from NY state COVID-19 patients uploaded to the SRA. Of the 7,309 samples publicly available as of July 21, 2021, none had sequences that matched the WNY lineages.

Arguing against the possibility of unsampled human strains is the geographical stratification of these cryptic lineages. Since January, the cryptic lineages have remained geographically constrained over many months in the sewersheds we sampled, which is not consistent with a contagious human pathogen. We suspect this lack of dispersal is consistent with infections of non-human animals with restricted movements or home ranges.

### Do Cryptic Lineages Indicate Presence of SARS-CoV-2 Animal Reservoirs?

Another hypothesis is that these lineages are derived from SARS-CoV-2 animal reservoirs. To date, there have been a number of animals infected by SARS-CoV-2, including in mink^25^, lions and tigers^26^, and cats and dogs^27–29^. To gain insight into the possible host range of these lineages, synthetic DNA coding for the amino acid sequences for these four lineages were generated and introduced into a SARS-CoV-2 Spike expression construct for functional analysis (Fig. 2). All four of these lineages were found to be fully functional and produced transduction-competent lentiviral pseudoviruses with titers similar to the parent strain (D614G). To determine if these pseudoviruses displayed an expanded receptor tropism, stable cell lines expressing Human, Mouse, or Rat ACE2 were cultured with the pseudoviruses (Fig. 2). While the parent SARS-CoV-2 Spike pseudoviruses could only transduce cells with Human ACE2, all four of the WNY lineages could efficiently transduce cells with the Human, Mouse, and Rat ACE2. Because some patient-derived SARS-CoV-2 lineages, such as Beta and Gamma, have also gained the ability to infect rodent cells (Fig. 2, N501Y+A570D), this observation cannot be taken as evidence that these lineages were derived from such a host^33^. Nonetheless, the observation is consistent with the possibility that these lineages are derived from an animal host such as a rodent.

**Figure 2.**
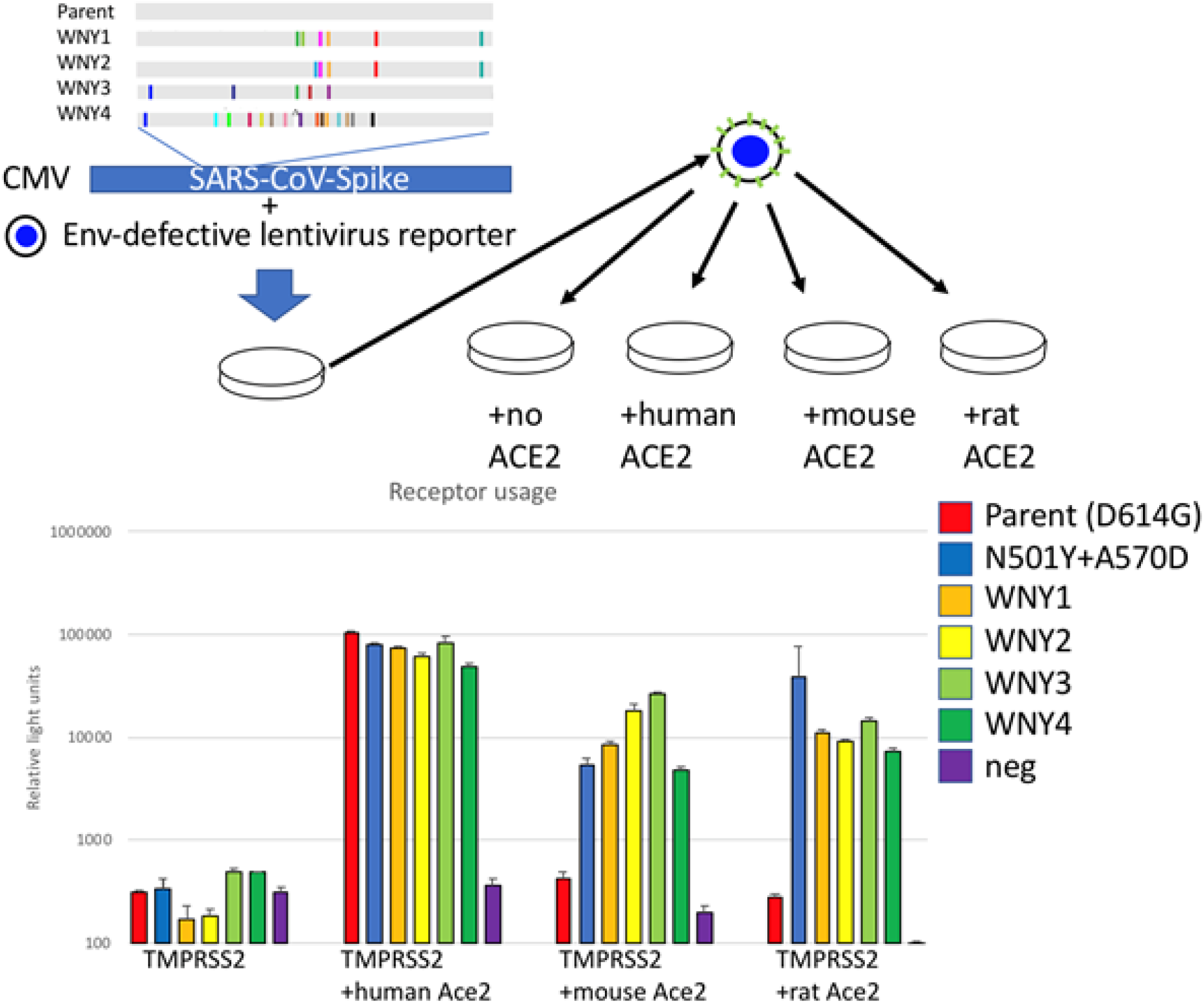
ACE2 usage by WNY lineages. A. Schematic of lineages and pseudovirion production. WNY1 = E484A/F486P/S494P/Q498Y/H519N/F572N, WNY2 = Q493K/ S494P/Q498Y/H519N/T572N, WNY3 = K417T/K444T/E484A/F590Y/Q498H, WNY4 = K417T/N439K/K444N/Y449R/L452R/N460K/S477N/Δ484/F486V/S494T/G496V/Q498Y/ N501T/G504D/505H/H519Q. Pseudovirions with indicated Spike proteins were generated and used to transduce 293FT+TMRPSS2 stably transduced with human, mouse or rat ACE2. Representative example of three experiments performed in triplicate.

If such reservoirs exist, the animal host would need to meet several criteria. First, the host species would likely need to be present in the sewershed. Second, the number of susceptible animals present must be high enough to sustain an epidemic for at least six months (i.e., the time period for which we observe these sequences). Third, host animals must not disperse beyond the geographical locations where the sequences are found. Finally, there must be a route for shed viruses to enter the sewersheds where the cryptic lineages are seen.

We considered several mammal species known to inhabit NYC that may meet these criteria, including bats (several species), cats (*Felis catus*), dogs (*Canis familiaris*), grey squirrels (*Sciurus carolinensis*), mice (*Mus musculus* or *Peromyscus leucopus*), opossums (*Didelphis virginiana*), rabbits (*Sylvilagus floridanus*), raccoons (*Procyon lotor*), rats (*Rattus norvegicus*), and skunks (*Mephitis mephitis*). To narrow our search, we reasoned that if viruses are being shed from one of these animals, then we should be able to detect rRNA from the animal in the sewershed as well.

### Mammalian Species Detected in Wastewater

RNA extracted from wastewater and amplified with 12S rRNA primers (Supplementary Table 2) was deep sequenced. We were able to detect mammalian rRNA in sewersheds where the cryptic lineages were found (Table 1). Several species, such as cow, pig, and sheep, are not indigenous to NYC. These detects are likely derived from food consumption so are ruled out as possible hosts. After non-indigenous mammals were removed, three remaining mammalian species were repeatedly detected: cats, dogs, and rats (Table 1). Cats and dogs are susceptible to SARS-CoV-2^30,31^, and cats are able to transmit to other animals^29^. Many rodents are not permissive for infection by the canonical SARS-CoV-2 strain^28,32^, but some variants have an expanded tropism that includes mice^33^. A 2013 census estimated that there are 576,000 pet cats in NYC households^34^, but this estimate does not include stray cats.

**Table 1.** Predominant species detected in NYC wastewater via deep sequencing of 12S amplicons. The results of two independent sequencing runs are shown. Data for a WWTP where no cryptic lineages were detected is included for comparison purposes (WWTP12).

| Species | Common Name | WWTP 3 | WWTP 10 | WWTP 11 | WWTP 12 |
| --- | --- | --- | --- | --- | --- |
| *Homo sapiens* | Human | 2/2 | 2/2 | 2/2 | 2/2 |
| *Bos taurus* | Cow | 2/2 | 2/2 | 2/2 | 2/2 |
| *Sus scrofa* | Pig | 2/2 | 2/2 | 2/2 | 1/2 |
| *Rattus norvegicus* | Rat | 2/2 | 0/2 | 2/2 | 2/2 |
| *Canis familiaris* | Dog | 1/2 | 2/2 | 2/2 | 1/2 |
| *Felis catus* | Cat | 1/2 | 1/2 | 2/2 | 1/2 |
| *Ovis aries* | Sheep | 1/2 | 0/2 | 0/2 | 0/2 |

Extrapolating from a limited study conducted in 2017 implies a stray cat population of about 2,500 animals^35^, but this number does not accord with the approximately 18,000 animals received annually by NYC Animal Care Centers^34^. There are currently 345,727 active dog licenses in NYC^36^, but this figure is likely a significant underestimate and the true number may be at least double this figure. Despite these uncertainties, both cat and dog populations are dwarfed by the NYC rat population, which is estimated to number between 2-8 million animals^37^.

WNY1 and WNY2 were both present in WWTP10. Cat and dog, but not rat, rRNA was observed in wastewater from this sewershed. We speculate that other species are the likely source of viruses shed into wastewater in this area. By contrast WNY3 and WNY4 were present in WWTPs 11 and 3 respectively where we consistently detected the presence of rat rRNA (Table 1).

### Lineages Detected from Wastewater Are Resistant to Some Neutralizing Antibodies

In addition to polymorphisms from the WNY lineages that are known to affect viral tropism, many of the polymorphisms are also known to affect antibody evasion. In particular, the WNY polymorphisms at positions K417, N439, N440, K444, L452, N460, E484, Q493, S494, and N501 have all been reported to evade neutralization by particular antibodies^4,38–41^. Most neutralizing antibodies against SARS-CoV-2 target the RBD of Spike, and most of these neutralizing antibodies are divided into 3 classes based on binding characteristics^42^.

To test if the WNY lineages have gained resistance to neutralizing antibodies, we obtained three clinically approved neutralizing monoclonal antibodies representing these 3 classes, LY-CoV016 (etesevimab, Class 1)^43^, LY-CoV555 (bamlanivimab, Class 2)^44^, and REGN10987 (imdevimab, Class 3)^45^, and tested their ability to neutralize the WNY lineages. All four of the WNY lineages displayed complete resistance to LY-CoV555, despite the parent lineage remaining potently sensitive to this antibody (Fig.3). The WNY 1 and 2 remained at least partially sensitive to LY-CoV016 and REGN10987, but WNY 3 and 4 appeared to be completely resistant to all three neutralizing antibodies (Fig. 3).

**Figure 3.**
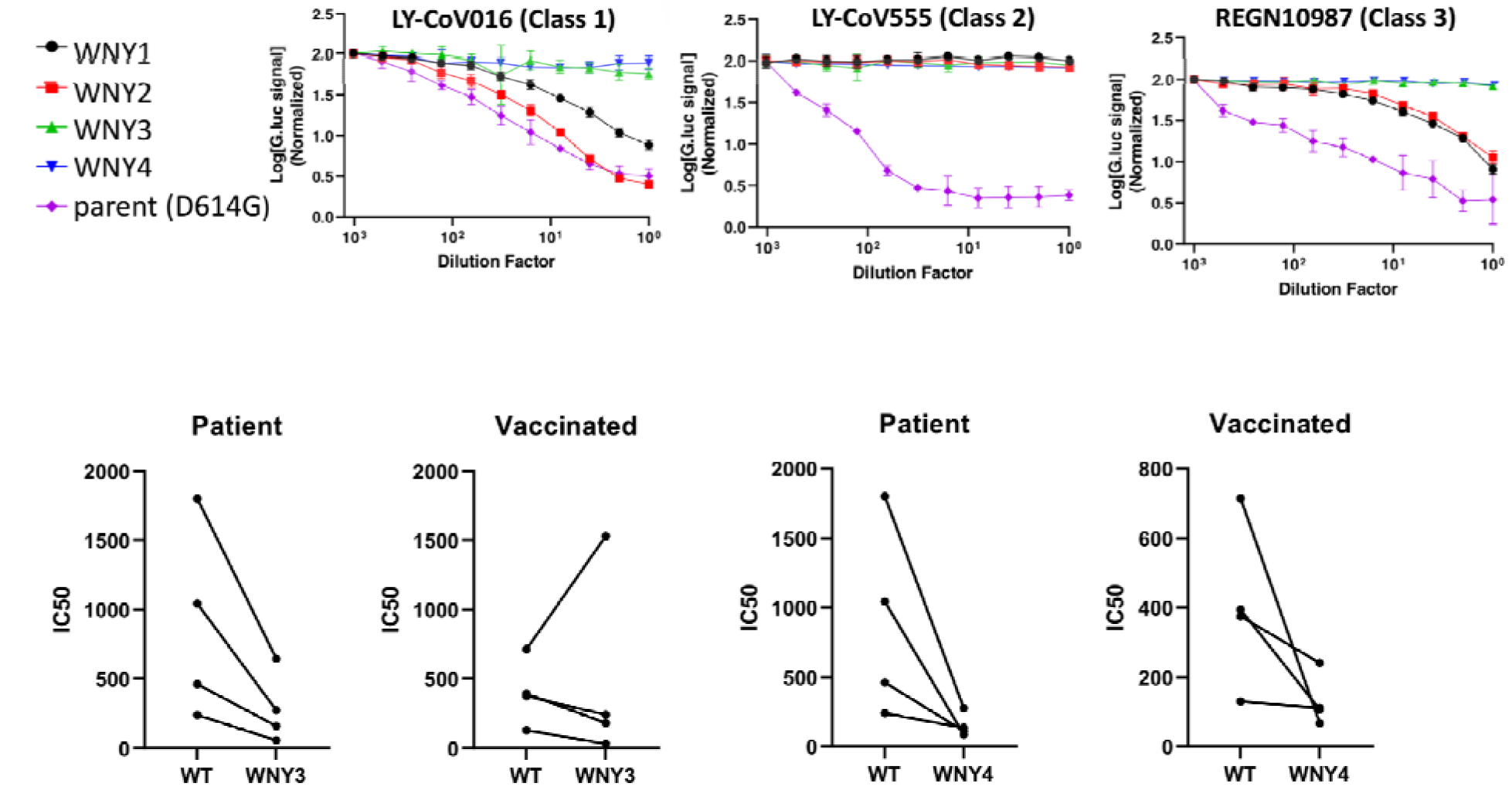
Antibody resistance to monoclonal neutralizing antibodies and patient plasma. Lentiviral reporter pseudoviruses containing parent (D614F), WNY1, 2, 3, or 4 Spike proteins were treated with 2-fold dilutions of indicated monoclonal neutralizing antibody and used to infect 293FT+TMPRSS2+human ACE2. Representative examples of 3 experiments performed in triplicate are shown.

Finally, we tested the ability of plasma from fully vaccinated individuals (Pfizer) or patients previously infected with SARS-CoV-2 to neutralize WNY 3 and 4. All patients’ plasma retained some capacity to neutralize these lineages (Fig. 3). However, previously infected patients had a greater reduction in ID50 (WT vs variant) than vaccinated patients and both were more affected by the WNY4 variant than the WNY 3. It must be noted that neutralizing antibody activity from vaccinated individuals is not solely directed against the Spike RBD. Therefore, if the full Spike proteins from these lineages with the additional mutations they carry were tested, the neutralization capacity against these lineages is likely to be even further diminished. Thus, the characteristics of these variant lineages provide them the capacity to be an increased threat to human health.

### Conclusions and Outlook

To date, most data on SARS-CoV-2 genetic diversity has come from the sequencing of clinical samples, but such studies may suffer limitations due to biases, costs and throughput. Here we demonstrate the circulation of several lineages of SARS-CoV-2 in the NYC metropolitan area that have not been detected by standard clinical surveillance. While the origins of these lineages have not been determined, we have demonstrated that these lineages have expanded receptor tropism which is consistent with expansion to an animal reservoir. Finally, we demonstrated that these lineages have gained significant resistance to some patient-derived neutralizing monoclonal antibodies. Thus, these novel lineages could be relevant to public health and necessitate further study.

## As a guideline, Methods sections typically do not exceed 3,000 words

### Wastewater Sample Processing and RNA Extraction

Wastewater was collected from the inflow at 14 NYC wastewater treatment plants and RNA isolated according to our previously published protocol^2^. Briefly, 250 mL from 24-hr composite raw sewage samples obtained from NYC WWTPs were centrifuged at 5,000 x g for 10 min at 4°C to pellet solids. 40 mL of supernatant was passed through a 0.22 μM filter (Millipore). Filtrate was stored at 4°C for 24 hrs after adding 0.9 g sodium chloride and 4.0 g PEG 8000 (Fisher Scientific) then centrifuged at 12,000 x g for 120 minutes at 4 °C to pellet the precipitate. The pellet was resuspended in 1.5 mL TRIzol (Fisher), and RNA was purified according to the manufacturer’s instructions.

### Targeted PCR

#### iSeq sequencing

RNA isolated from wastewater was used to generate cDNA using ProtoScript® II Reverse Transcriptase (New England Biolabs). The RNA was incubated with an RBD specific primer (ccagatgattttacaggctgcg) and dNTPs (0.5 mM final concentration) at 65°C for 5 minutes and placed on ice. The RT buffer, DTT (0.01 M final concentration), and the RT were added to the same tube and incubated at 42°C for 2 hours followed by 20 minutes at 65°C to inactivate the enzyme. The RBD region was amplified using Q5^®^ High-Fidelity DNA Polymerase using primers that incorporate Illumina adaptors. PCR performed as follows: 98°C(0:30) + 40 cycles of [98°C(0:05) + 53°C(0:15) + 65°C(1:00)] x 40 cycles + 65°C (1:00).

The RBD amplicons were purified using AMPure XP beads (Beckman Coulter). Index PCR was performed using the Nextera DNA CD Indexes kit (Illumina) with 2X KAPA HiFi HotStart ReadyMix (Roche), and indexed PCR products purified using AMPure beads. The indexed libraries were quantified using the Qubit 3.0 and Qubit dsDNA HS Assay Kit and diluted in 10 mM Tris-HCl to a final concentration of approximately 0.3 ng/μL (1 nM). The libraries were pooled together and diluted to a final concentration of 50 pM. Before sequencing on an Illumina iSeq100, a 10% spike-in of 50 pM PhiX control v3 (Illumina) was added to the pooled library.

#### MiSeq sequencing

The primary RBD RT-PCR was performed using the Superscript IV One-Step RT-PCR System (Thermo Fisher Scientific).□Primary RT-PCR amplification was performed as follows: 25°C(2:00) + 50°C(20:00) + 95°C(2:00) + [95°C(0:15) + 55°C(0:30) + 72°C(1:00)] x 25 cycles using the MiSeq primary PCR primers (Table 1). rRNA amplification used the same primary reaction conditions except containing 30 cycles using previously described 12s^46^ and 16s primers^47^. Secondary PCR (25 µl) was performed on RBD amplifications using 5 ul of the primary PCR as template with MiSeq nested gene specific primers containing 5’ adapter sequences (Table 1) (0.5 µM each), dNTPs (100 µM each) and Q5 DNA polymerase (New England Biolabs).□Secondary PCR amplification was performed as follows: 95°C(2:00) + [95°C(0:15) + 55°C(0:30) + 72°C(1:00)] x 20 cycles.□A tertiary PCR (50 µl) was performed to add adapter sequences required for Illumina cluster generation with forward and reverse primers (0.2 µM each), dNTPs (200 µM each), and Phusion High-Fidelity DNA Polymerase (1U) (New England Biolabs).□PCR amplification was performed as follows: 98°C(3:00) + [98°C(0:15) + 50°C(0:30) + 72°C(0:30)] x 7 cycles +72°C(7:00).□Amplified product (10 µl) from each PCR reaction is combined and thoroughly mixed to make a single pool.LPooled amplicons were purified by addition of Axygen AxyPrep MagPCR Clean-up beads in a 1.0 ratio to purify final amplicons.□The final amplicon library pool was evaluated using the Agilent Fragment Analyzer automated electrophoresis system, quantified using the Qubit HS dsDNA assay (Invitrogen), and diluted according to Illumina’s standard protocol.□The Illumina MiSeq instrument was used to generate paired-end 300 base pair length reads. Adapter sequences were trimmed from output sequences using cutadapt.

### Wastewater rRNA Sequencing

cDNA from wastewater was also used to generate libraries using the primers indicated in Table 1. rRNA Libraries were amplified using ProtoScript^®^ II Reverse Transcriptase (New England Biolabs) and pooled and sequenced on the iSeq100 and MiSeq as described above.

### Bioinformatics

iSeq reads were uploaded to the BaseSpace Sequence Hub, and demultiplexed using a FASTQ generation script. Reads were processed using the published Geneious workflows for preprocessing of NGS reads and assembly of SARS-CoV-2 amplicons^48^. Paired reads were trimmed, and the adapter sequences removed with the BBDuk plugin. Trimmed reads were aligned to the SARS-CoV-2 reference genome MN908947. Variants present at frequencies of 1% or above were called using the Annotate and Predict Find Variations/SNPs in Geneious and verified by using the V-PIPE SARS-CoV-2 application^49^.

Reads from iSeq and MiSeq sequencing were processed as previously described^6^. Briefly, VSEARCH tools were used to merge paired reads and dereplicate sequences^50^. Dereplicated sequences from RBD amplicons were respectively mapped to the reference sequence of SARS-CoV-2 (NC_045512.2) Spike ORF using either Bowtie2^51^ or Minimap2^52^. Mapped RBD amplicon sequences were then processed with SAM Refiner using the same Spike sequence as a reference and the command line parameters ‘--alpha 1.8 --foldab 0.6’. The output from SAM Refiner were reviewed to determine the known and novel lineage makeup of the sampled sewersheds.

For sequencing from rRNA templates, dereplicated reads were mapped with Bowtie2 and Minimap2 to a collected reference index of mitochondrial and rRNA related animal sequences from NCBI’s nucleotide and refseq databases (https://www.ncbi.nlm.nih.gov/). Mapped rRNA sequences were reviewed for matching of specific organisms. Sequences with poor mapping to sequences in the index and a random selection of sequences with good mapping were checked by Blast (https://blast.ncbi.nlm.nih.gov/Blast.cgi) to verify the organism match. Matches were corrected based on the blast results as needed.

For both iSeq and MiSeq datasets, we examined Outbreak.info for the prevalence of each mutation and their associated lineages in New York, the United States and worldwide (Supplemental Table 1).

For sequences from GISAID, fasta formatted sequences were obtained from the GISAID database for submissions from NYC patients from January to April, 2021. These sequences were processed similarly to the dereplicated sequences above. Minimap2 was used to map the sequences to the Spike ORF, then SAM Refiner was used to process the mapped sequences using ‘--min_count 1 --min_samp_abund 0’ parameters to include all variations in the output.

For SRA data, SRAs were initially searched using NCBI SRA Blast for “TTTAAAATCATATGGTTTCTAT”, the sequence encoding Q493K and Q498Y of SARS-CoV-2 Spike. Also, fastq formatted sequences were obtained for sequencing runs from NY state SARS-CoV-2 clinical samples and all SARS-CoV-2 wastewater samples as of 7-21-2021. Fastq files were processed similarly to MiSeq data, with the merging step skipped for unpaired reads. Reads mapped to the Spike Orf were processed with SAM Refiner with the parameters ‘--wgs 1 --min_count 1 -- min_samp_abund 0’.

### Plasmids

Eukaryotic expression vectors for the heavy and light chains of antibodies LY-CoV016, LY-CoV555, and REGN10987 were obtained from Genscript. The lentiviral reporter constructed containing *Gaussia* luciferase (Gluc) with a reverse-intron (HIV-1-GLuc) was previously described^53^. The codon-optimized SARS-CoV-2 Spike expression vector was obtained from Tom Gallagher^54^. This construct was modified to enhance transduction efficiency by truncating the last 19 amino acids, and introducing the D614G amino acid change. DNA gBlocks containing the WNY RBD sequences were synthesized by IDT and introduced into the SARS-CoV-2 expression construct using In-Fusion cloning (Takara Bio). Lentiviral Mouse and Rat Ace2 vectors pscALPSpuro-MmACE2 (Mouse) and pscALPSpuro-RnACE2 (Rat) were obtained from Jeremy Luban^55^.

### Cell culture

The 293FT cell line was obtained from Invitrogen. The 293FT+TMPRSS2 and 293FT+TMPRSS2+human Ace2 cells were previously described^56^. All cells were maintained in Dulbecco’s modified Eagle’s medium (DMEM) supplemented with 10% fetal bovine serum, 2mM L-glutamine, 1 mM sodium pyruvate, 10 mM nonessential amino acids, and 1% minimal essential medium (MEM) vitamins. The ACE2 cell lines were generated by transfecting 293FT cells with 500 ng HIV GagPol expression vector, 400 ng of pscALPSpuro-MmACE2 (Mouse) or pscALPSpuro-RnACE2 (Rat), and 100 ng of VSV-G expression vector. Viral medium was used to transduce 293FT+TMPRSS2 cells^56^, and cells were selected with puromycin (1 mg/mL) beginning 2 days postransduction and were maintained until control treated cells were all eliminated.

### Monoclonal antibody synthesis

Transfections of 10cm dishes of 293FT cells were performed 5 mg each of heavy and light chain vectors and 40 mg polyethyleneimine (PEI)^57^.

### Virus production and infectivity assays

All transfections were performed in 10cm dishes. 293FT cells were transfected with a total of 9 mg of HIV-1-Gluc, 1 mg of CMV Spike vector, and 40 mg of PEI^57^. Supernatants containing the virus were collected 2 days post-transfection. Transduction of ACE2 expressing cells was performed by plating 30,000 cells in 96 well plates and co-culturing with 50 mL of HIV-1-GLuc/Spike particles. Gluc was measured 2 days post-transduction.

### Antibody Neutralization Assay

All blood collection and processing were performed under the approved protocols (MU Study of Serology for SARS-CoV-2 and MU COVID19 Vaccine study) by the Institutional Review Board of the University of Missouri. Written consent was received from all human subjects prior to being enrolled in the study. Subjects were requested to provide a date of positive PCR test for SARS-CoV-2 and subsequently had laboratory-based serologic tests to confirm the presence of antibody against SARS-CoV-2 S1 RBD protein. A total of 10-20 mL of blood was collected from each participant. The plasma was then separated from the blood cells by centrifugation and stored at -80°C.

### Pseudovirus Neutralization Assay

All human plasma samples were heat inactivated for 30 min at 56°C prior to the assay. Samples were diluted at 2-fold in 10 serial dilution in duplicates. Serially diluted samples were incubated with pre-titrated amounts of indicated pseudovirus at 37°C for 1 hour before addition of 293FT cells expressing human ACE2 and TMPRSS2 at 30,000 cells per well. Cells were incubated for 2 days and then the supernatant was used to measure gaussian luciferase (RLU). Infection was normalized to the wells infected with pseudovirus alone. Neutralization IC50 titers were calculated using nonlinear regression (Inhibitor vs normalized response—variable slope) in GraphPad Prism 9.0.

## Data Availability

All sequencing raw reads available in NCBI'S SRA

https://docs.google.com/spreadsheets/d/1HXgCFhTFO2UxTQQaXgtxdWbJdmr2fXbd7mO3RzV_sjc/edit?usp=sharing

## Data Availability

Raw sequencing reads are available in NCBI’s Sequence Read Archive (SRA) under accession # PRJNA715712.

## Acknowledgments

The research described herein would not be possible if not for the assistance and support of a wide-range of organizations and individuals that came together to address the shared calamity that is the COVID-19 pandemic. We thank Jasmijn Baaijens, Michael Baym, Gina Behnke, Esmeraldo Castro, Francoise Chauvin, Alexander Clare, Pilar Domingo-Calap, Robert Corrigan, Pam Elardo, Raul Gonzalez, Crystal Hepp, Catherine Hoar, Dimitrios Katehis, William Kelly, Samantha McBride, Hope McGibbon, Hilary Millar, Jason Munshi-South, Samantha Patinella, Krish Ramalingam, Andrea Silverman, Jasmin Torres, Arvind Varsani, Peter Williamsen, and members of the Dennehy Lab for for support, advice, discussions, and feedback. We also thank Molly Metz for assistance with graphics and figure design. This work was funded in part by the New York City Department of Environmental Protection, a donation from the Linda Markeloff Charitable Fund, and from the National Institutes of Health grant U01DA053893-01. The Water Research Foundation, the NSF Research Coordination Network for Wastewater Surveillance for SARS-CoV-2 and Qiagen Inc. provided resources, materials and supplies, technical support, and community support. Special thanks to Vincent Racaniello and the team at This Week in Virology for connecting the New York and Missouri teams.

## Author Information

### Contributions

M.T., D.S.S., M.C.J., M.D. and J.J.D. supervised the project. M.T., D.S.S., M.C.J., and J.J.D. conceptualized the project. M.T., S.K., D.S.S., M.C.J., MD, and J.J.D. designed experiments. D.S.S., M.T., K.C., A.G., S.K., N.K., K.M.S., G.S., M.G., R.S., C.R., Y.G. and F.S. performed experiments. D.S.S., D.G., I.H., M.M., N.M., M.C.J., D.A.G. T.D.L. and J.J.D. performed data analysis and interpretation. M.T., D.S.S., D.A.G., M.C.J. and J.J.D. wrote the original and revised manuscript drafts. All authors contributed to reviewing and editing of the manuscript.

## Ethics Declarations

The authors declare no competing financial interests.

## Additional Information

Supplementary Information is available for this paper.

Correspondence and requests for materials should be addressed to JJD or MCJ.

---

**Supplementary Table 1. Mutations observed in NYC wastewater**

**https://docs.google.com/spreadsheets/d/1HXgCFhTFO2UxTQQaXgtxdWbJdmr2fXbd7mO3RzV_sjc**

**Supplementary Table 2.**
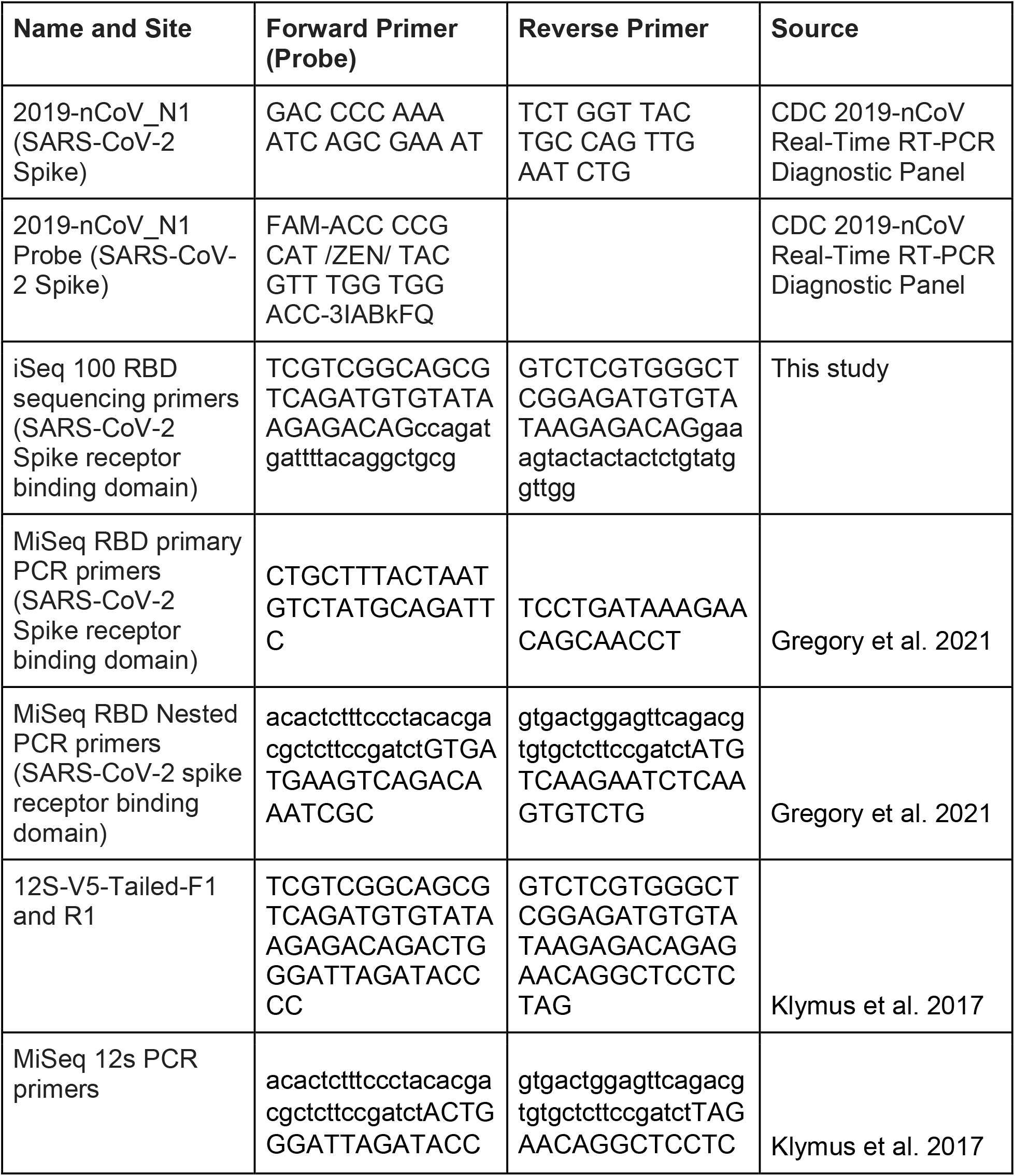
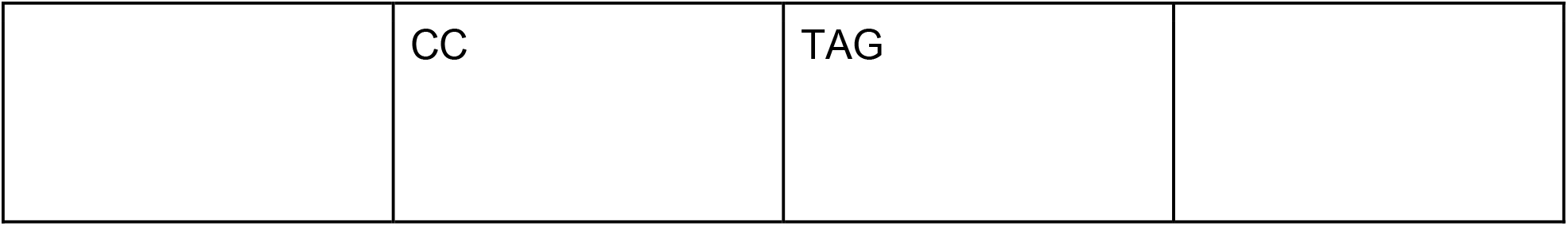
Primers and probes used in this study.

## Notes

### Competing Interest Statement

The authors have declared no competing interest.

### Author Declarations

Approved by Queens College IRB

